# Efficacy and Safety of Polyherbal formulation as an add-on to the standard of care in mild to moderate COVID-19: A randomized, double-blind, placebo-controlled trial

**DOI:** 10.1101/2021.05.14.21256900

**Authors:** Suresh B. Patankar, Hrishikesh Rangnekar, Kalpana Joshi, Kishor Suryawanshi, Pravin Soni, Anupama Gorde, Tejas Shah, Sagar Patankar, Diwakar Jha, Rajesh Raje

## Abstract

**Objective:** To assess the efficacy and safety of polyherbal formulation (designated as IP) in comparison to placebo as add on to the standard of care (SoC) among patients with mild to moderate novel corona virus disease 2019 (COVID-19)

**Methods:** Laboratory proved patients of mild to moderate COVID-19 disease were randomized to either placebo or IP as an add-on to SoC. Using quantitative reverse transcription-polymerase chain reaction (qRTPCR), we assessed the effect on viral load (VL). Change in immunological parameters such as blood lymphocyte subset, serum immunoglobulin was determined. The clinical improvement was assessed using a numerical rating scale (NRS) and WHO ordinal scale. Patients were followed for 30 days after randomization.

**Results:** In total, 72 patients were randomized to either placebo (n=33) and IP (n=39). Fifty-two patients (n=21 in placebo and n=31 in IP arm) had qRT-PCR on day 0 and day 4. There was significant reduction in VL in IP arm (from 662081 copies/mL on day 0 to 48963 copies/mL on day 4; *p=*0.002)) but not in the placebo arm (from 385670 copies/mL on day 0 to 66845 copies/mL on day 4, *p=*0.106). Change in the NRS score and WHO ordinal scale score was significant in both treatment arms. However, the difference between the two groups was statistically significant in favour of drug group,. The increase in Th1 response was significant in the IP arm (p=0.023) but not in the placebo arm (p=0.098), thus implying immunomodulatory activity in the drug. No safety concerns were observed in any of the trial participants.

**Conclusion:** This study finds that polyherbal formulation reduces viral load and contributes to immunomodulation and improvement in clinical conditions when used as add-on to the standard care in patients with mild to moderate COVID-19 without any side effects. These findings need to be further confirmed in a large, prospective, randomized study.

## INTRODUCTION

Novel coronavirus disease 2019 (COVID-19) pandemic is a significant contributor to morbidity and mortality in affected individuals. World Health Organization, as of 23April 2021, reports 144,358,956 confirmed cases of COVID-19, including 3,066,113 deaths [1]. Since its first identification in December 2019, there is now a fair understanding of the COVID-19 disease pathogenesis. Hyper stimulated immune response causing systemic inflammatory response syndrome or “cytokine storm” is known to be implicated in progression to severe COVID-19 [2,3]. Such immune response correlates linearly with the viral load that in turn correlates with disease severity [4,5]. Modulating the immune response in COVID-19 is now an established treatment approach. The RECOVERY trial showed that dexamethasone 6 mg once daily for ten days significantly lowered 28-day mortality in hospitalized COVID-19 patients [6]. Also, treatment with specific interleukin -6 inhibitor tocilizumab did not improve survival but reduced the likelihood of progression to the composite outcome of mechanical ventilation or death [7]. With Itolizumab, an anti-CD6 humanized monoclonal antibody with an immunomodulating action on Teffector cells, there was a significant reduction in 30-day mortality along with significant improvement in clinical immunological, and oxygen parameters [8]. Thus, modulating immune response can help to improve clinical parameters and mortality outcomes, especially in hospitalized moderate to severe COVID-19 patients.

Herbal formulations have long been assessed for their potential immune modulating effects [9,10]. In an in-vitro study (*data on file*), we observed that a combination of herbals (IP) such as Ashwagandha (*Withaniasomnifera)*, Vidanga (*Embeliaribes*), Guduchi (*Tinospora cordifolia)*, Haritaki (*Terminalia chebula*), Aamalaki (*Emblica officinalis*), Shatavari (*Asparagus racemosus*), Yashtimadhu (*Glycyrrhiza glabra*), Ginger (*Zinziber officinale*), Pippali (*Piper longum*), Shankha Bhasma and Jasath Bhasma showed antiviral activity against SARS-CoV-2. As the search for potential antiviral and immune modulating therapy for COVID-19 is ongoing, this could be potentially beneficial. Therefore, we conducted this randomized, double-blind, placebo-controlled study to determine the effect of the polyherbal formulation on viral load and immunological parameters and clinical improvement in mild to moderate COVID-19 disease when used as an add-on to the SoC.

## MATERIALS AND METHODS

### Design and Population

This study was a double-blind, randomized, placebo-controlled trial to assess the safety and efficacy of polyherbal drug formulation (designated as IP) in patients with mild to moderate COVID-19. The study was conducted at a single center in Pune, Maharashtra, India. We enrolled adults of age 18 years and above, diagnosed with laboratory-proven COVID-19 disease of mild to moderate severity who were admitted to the hospital as per the isolation protocol. Pregnant and lactating females, any respiratory symptoms of >7 days, patients with known pathology affecting the respiratory system, diagnosed hematological disorders, patients with a terminal illness, any other condition which in the view of the investigator would interfere with the general clinical well-being of the participant and those not willing to participate in the study were excluded. The trial was conducted as per the ethical principles of the Declaration of Helsinki, good clinical practice recommendations and applicable local regulatory guidelines. The study was approved by the Board of Research Studies (BORS) and Institutional Ethics Committee of Yashwantrao Chavan Memorial Hospital, Pimpri, Pune. Informed consent was obtained from all the participants before enrolment. The study was registered with the clinical trial registry of India (CTRI/2020/07/026570).The study protocol was published in a peer-reviewed journal [11].

### Study treatments and follow-ups

Patients with a known positive status of RT-PCR for SARS-CoV-2 disease were screened for the study. Eligible and consenting patients were enrolled. Patients were randomized using computer-generated randomization codes to receive either IP or placebo as an add-on to the recommended standard care (SoC) for COVID-19. IP in the title was a combination of IP1 and IP2, having the following ingredients. IP1: Shunthi (*Zinziber officinale*), Vidanga (*Embelia ribes*), Yashtimadhu (*Glycyrrhiza glabra*), Shankhabhasma and Jasath Bhasma. IP2 contains: Haritaki (*Terminalia chebula*), Guduchi (*Tinospora cordifolia)*, Shatavari (*Asparagus racemosus*), Aamalaki (*Emblica officinalis*), Pippali (*Piper longum*), Ashwagandha (*Withania somnifera)* and Jasath Bhasma. The investigational products were manufactured per API guidelines. Patients were advised to take one capsule, two times a day (of each IP1 and IP2) or placebo, after meals. Patients received a prepacked carton of study treatment for a total duration of 30 days. Capsule IP1 and IP2 were given for the first 15 days, and last 15 days they were given only IP2. Patients receiving placebo also received 2 capsules twice a day for first 15 days and for next 15 days, one capsule twice a day (Image 1). All the patients continued their routine diet and physical activity and other prescribed treatment as per Indian Council of Medical Research (ICMR) and World Health Organization (WHO) guidelines in circulation at the time of study conduct.

During baseline visit (Day 0) nasopharyngeal swab was taken for qRTPCR, numerical rating scale (NRS) was filled for clinical status and the blood was withdrawn for various immunological inflammatory and safety markers. Patients had three follow-ups during the 30-day treatment period. First follow-up visit was done at day 4±1. qRTPCR, NRS was repeated at this visit and any clinical adverse events were recorded. Second follow-up was done clinically or telephonically (if they were discharged) on day 15±2 to record any clinical deterioration in clinical condition and to record adverse effects if any. Last follow-up was at day 30±2 to document clinical condition, adverse effects and perform laboratory investigations. In this last follow-up, study personnel visited patients in their homes to avoid their repeat visit to site as it was active COVID-19 hospital. Patients were asked to visit any time if they observed any deterioration in clinical condition or had developed any untoward reactions.

#### Quantitative estimation of viral load using qRTPCR

Three viral loci RdRP, N gene and E gene and a human gene as internal control (RNaseP) were detected using GenePath Diagnostics’ indigenously developed qRTPCR assay, CoViDx One v2.1.1 keeping appropriate controls and standards. Ten micro litres of extracted nucleic acid was used as an input for the qRT-PCR. Quantitative estimation for each genomic target was performed using standards, for which the number of genome copies were already known. The threshold cycle (Ct value) for each genomic target was extrapolated to the viral load of standards to estimate the number of copies/uL which were then converted to copies/mL for each gene for each sample. Average number of copies/ ml was calculated for each sample that accounts for the approximate viral load in the sample. Ct values for control human gene (RNaseP) ensured the adequacy of sample and served as control for nucleic acid extraction and nucleic acid amplification.

On the day of first encounter with patients, they were subjected to thorough clinical and laboratory investigations. After signing of informed consent, their demographic details, and clinical parameters were recorded. Oxygen saturation was assessed using pulse oximeter. We investigated viral load (VL) with quantitative RTPCR for COVID-19, and blood was drawn for lymphocyte subset analysis (absolute neutrophil count, absolute lymphocyte count, TH1, TH2, Treg cells, NK Cells and CD markers such as CD3, CD4, CD3: CD8 ratio) as well as serum IgG and IgM levels (not specific to COVID-19). Liver function tests and renal function tests were performed as a part of safety assessment.. Blood investigations were repeated on day 30 for immunological markers, and for safety assessments. We performed all the tests at a single accredited laboratory.

### Outcome measures

Primary efficacy outcome measure was change in viral load. Secondary efficacy outcome measures were change in disease severity score and change in immunological and inflammatory markers. Numerical Rating Scale (NRS) was used for overall disease severity. 0 was considered for normal, 1 to 3 for mild, 4 to 6 for moderate and 7 to 10 for severe severity of the disease. Patients were asked to rate the disease severity on the NRS. Also, change in immunological parameters such as lymphocyte subset analysis (TH1, TH2, Th17),NK Cells and CD markers, serum IgG and IgM levels and Covid19 antibodies were assessed. Inflammatory and other markers like C - reactive protein and D-Dimer, Sr. Ferritin were assessed in a small portion of enrolled subjects. Primary safety measure was incidence of adverse reactions (clinical and/or laboratory parameters).

## Statistical analysis

Sample size:

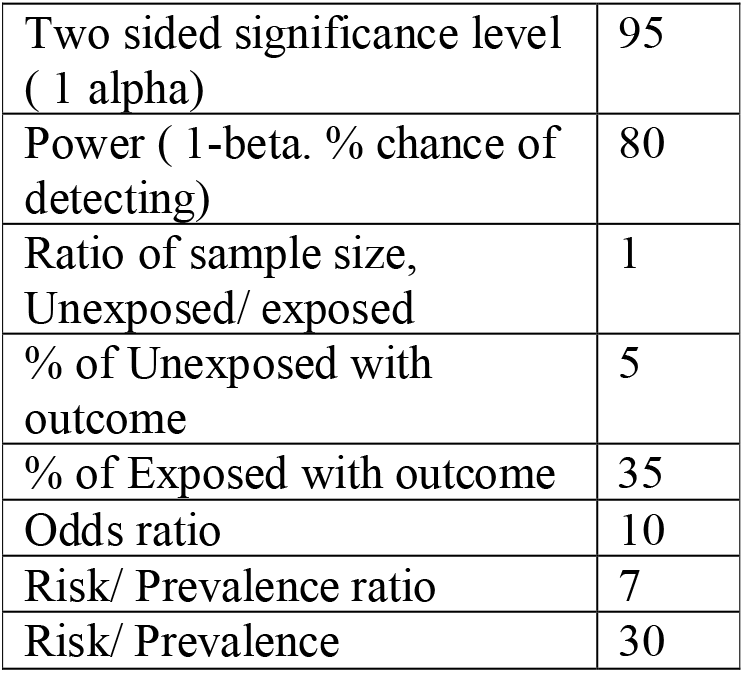

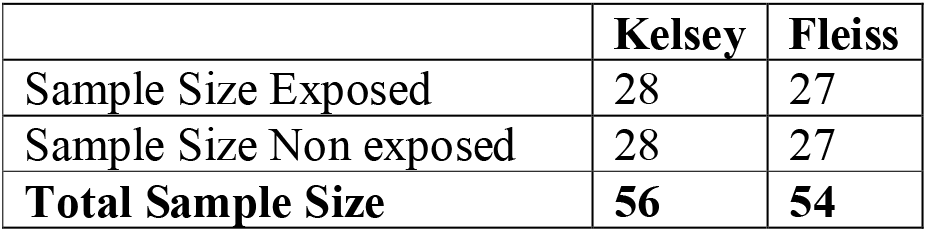

## Supporting information

Protocol--EC Approved

Consort Checklist

ETHICS STATEMENT

## Data Availability

Data cannot be shared publicly because the organization is a semi government
institution and access to the clinical records is restricted. This data are available from
the Institutional Data Access upon requesting Dr. Pravin Soni(M.D (Gen.Med), HoD Dept
of Med.) for researchers who meet the criteria
for access to confidential data.
Data on investigations like viral load and few haematological investigations can be
made available without any restrictions.

https://doi.org/10.1186/s13063-020-04906-x

## Results are rounded up to the nearest integer Results from OpenEpi, Version 3, open source calculator –SSC cohort

Considering drop-outs, 72 patients were enrolled. Data was entered in to the Microsoft excel sheet version 2016 and was analyzed with SPSS software. Categorical variables were presented as frequency and percentage. Statistical comparisons of categorical data were done with Chi square test or Fischer exact test. For continuous variables, normal distribution was assessed by plotting the histograms. Normally distributed data was presented using mean and standard deviations (SD) and non-normally distributed data presented as median (Interquartile range (IQR) 25-75). Student t-test and Mann-Whitney U test were applied accordingly to test the statistical differences in continuous independent variables. For before and after analysis, we used paired t test and Wilcoxon signed rank sum test to determine the statistical significance. P value <0.05 was considered significant for all comparisons.

## RESULTS

### Participant enrolment flow

Between August and September 2020, we screened 92 patients of which 72 were randomized to either placebo (n=33) or IP (n=39). Figure 1 shows study flow of the participant. Overall, 66 patients completed the 30-day follow-up (placebo n= 31; IP n=35).

**Figure 1:**
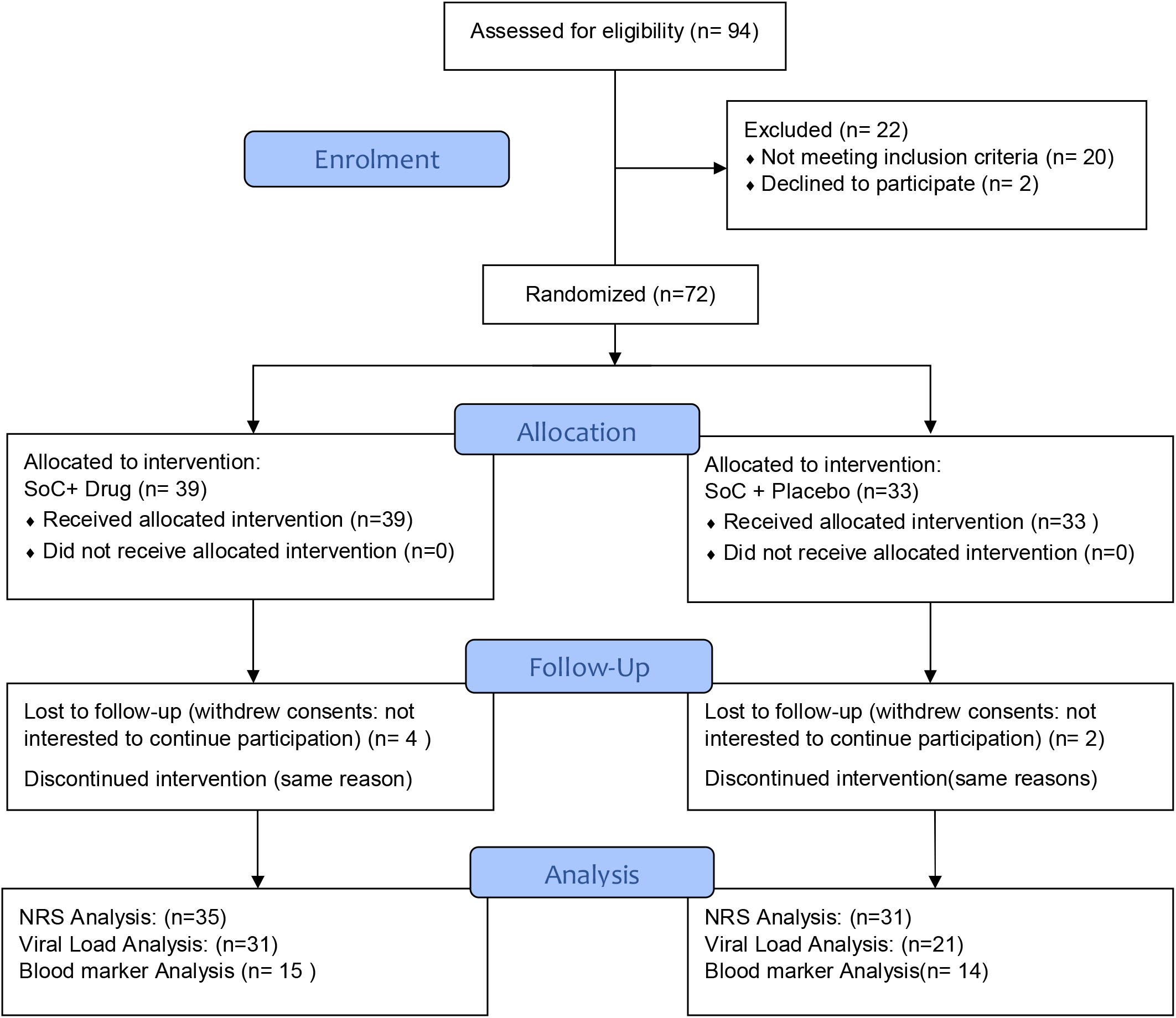
Consort Study flow Chart.

### Demographic parameters (Table no. 1)

**Table 1:** Baseline characteristics in two groups.

| Parameters | SoC + Placebo (n=33) | SoC + IP (n=39) | P value<br>(between groups) |
| --- | --- | --- | --- |
| Age (years) | 43.0 (34.0 - 58.0) | 47.0 (40.0 – 56.0) | 0.336 |
| Gender |  |  |  |
| Male | 20 (60.6) | 17 (43.6) | 0.150 |
| Female | 13 (39.4) | 22 (56.4) |  |

Data presented as median (IQR) or n (%); \*SoC = Standard of Care

Mean age was 43 years and 47 years in placebo and IP group (p=0.336). There was no difference in proportion of males and females in two groups (p=0.150).

### Effect on Viral Load (VL) (Table no. 2)

**Table 2:** Change in viral load in two groups.

| Viral load | SoC + Placebo<br>(N=21) | SoC + IP<br>(N=31) | P value<br>(between groups) |
| --- | --- | --- | --- |
| Baseline | 385670<br>(30807 to 131511846) | 662081<br>(56904 to 19485712) | 0.758 |
| Day 4 | 66845<br>(11978 to 836124.5) | 48963<br>(8774 to 1248257) | 0.845 |
| P value | 0.106 | 0.002 |  |

Overall, 52 patients had undergone qRT-PCR on day 4 (31 in drug arm, 21 in placebo arm). IP treatment was associated with significant reduction in median VL (from 662081 virus copies/mL (IQR: 56904 to 19485712) on day 0 to 48963 copies/mL (IQR: 8774 to 1248257) day 4; *p=*0.002)). In placebo group also, there was reduction in VL but it was non-significant (from 385670 copies/mL (IQR: 30807 to 131511846) on day 0 to 66845 copies/mL (IQR: 11978 to 836124.5) on day 4 (*p=*0.106)). Standard of care was continued in both the groups.

### Clinical Assessment (Table No. 3)

**Table 3:** Clinical Improvement.

| NRS score | SoC + Placebo (n=31) | SoC + IP (n=35) | P value<br>(between groups) |
| --- | --- | --- | --- |
| Baseline | 4.26±1.09 | 4.3±1.13 | 0.781 |
| Follow-up | 3.16±1.03 | 1.74±1.03 | <0.0001 |
| P value | <0.0001 | <0.0001 |  |
| **WHO Ordinal Scale** | **SoC + Placebo (n=31)** | **SoC + IP (n=35)** |  |
| Baseline | 1.71 ($\pm 0.78$) | 1.63 ($\pm 0.84$) | 0.58 |
| Follow-up | 1.26 ($\pm 0.63$) | 1.03 ($\pm 0.77$) | 0.33 |
| P value | 0.03 | 0.0001 |  |

In the IP arm, there was significant reduction in NRS from day 0 to day 4 (4.3 (+1.13) 1.74+(1.03), p<0.0001). In Placebo arm as well, there was significant reduction in NRS score (4.26+1.09 to 3.16±1.03, p<0.0001). Baseline scores of both the groups were similar, with P value 0.78; the difference between drug and placebo arm of ‘after’ was statistically highly significant. Later, upon WHO ordinal scale was available, and retrospectively the scores were assigned to the patients’ condition, on day 0 and 4, IP arm was at 1.63 (±0.84) and 1.03 (±0.45) respectively with p value 0.0001; whereas, for placebo, these values were 1.71 (±0.78)and 1.26 (±0.63) with p value 0.03. P value for drug arm was highly significant (>0.01).

### Blood Biomarkers for immunity and inflammation

Changes in the inflammatory and immunological parameters are shown in Table4, between day 0 and day 30. There was significant increase in absolute B cell count, absolute T cell count, absolute CD3, CD4 and T helper cells as well as in the ratio of absolute CD3 and CD8 cells in both IP and placebo arm. Change in absolute NK cell count was significant in placebo arm (p=0.020) but not in drug arm (p=0.067). However, the magnitude of change was higher with IP than the placebo. The increase in Th1 response was significant with IP treatment (p=0.023) but not with the placebo treatment (p=0.098). The IgG antibodies reduced significantly in both placebo and IP arm. However, the effect on IgM was non-significant in both groups. Covid antibodies were done on day 30. P value was not significant, however around 20% more antibodies were noted in drug arm than placebo arm. Due to inadequate sample size for CRP, D Dimer analysis was not feasible, however, average 50% reduction in CRP was seen in SoC + drug arm whereas increase in the SoC + Placebo arm was seen. Similar trend was seen for D Dimer also. Less number of patients opted for blood withdrawal during follow-up visits, and majority of them denied the blood letting.

**Table 4:** Changes in inflammatory and immunological parameters.

| Parameters | SoC + Placebo (n=14) | SoC + IP (n=15) | P value (between groups) |
| --- | --- | --- | --- |
| **Total leucocyte count** |  |  |  |
| Baseline | 6257.1±4488.6 | 7240.0±2580.4 | 0.472 |
| Follow-up | 7200.0±1669.6 | 7606.7±1547.5 | 0.502 |
| Change from baseline | 942.6 (3005.0 to -1119.3) | 366.7 (1720.2 to -986.8) |  |
| P value | 0.341 | 0.570 |  |
| **Absolute B cell count** |  |  |  |
| Baseline | 193.4±102.6 | 255.9±154.7 | 0.215 |
| Follow-up | 317.9±213.5 | 437.3±270.5 | 0.200 |
| Change from baseline | 124.5 (205.1 to 43.8) | 181.4 (282.4 to 80.5) |  |
| P value | 0.005 | 0.002 |  |
| **Absolute T cell count** |  |  |  |
| Baseline | 710.5±389.4 | 836.8±455.6 | 0.431 |
| Follow-up | 1512.7±477.8 | 1720.9±611.7 | 0.319 |
| Change from baseline | 802.2 (1094.5 to 510.0) | 884.1 (1230.7 to 537.5) |  |
| P value | <0.0001 | <0.0001 |  |
| **Absolute CD3, CD4 and T helper cells** |  |  |  |
| Baseline | 399.8±239.1 | 515.5±314.8 | 0.278 |
| Follow-up | 834.8±261.3 | 1055.7±417.6 | 0.102 |
| Change from baseline | 435.0 (628.1 to 242.1) | 540.2 (737.1 to 343.4) |  |
| P value | <0.0001 | <0.0001 |  |
| **Absolute CD3: CD8** |  |  |  |
| Baseline | 270.7±128.6 | 299.7±183.5 | 0.629 |
| Follow-up | 571.4±195.7 | 607.3±258.8 | 0.679 |
| Change from baseline | 300.7 (406.1 to 195.4) | 307.6 (455.8 to 159.3) |  |
| P value | <0.0001 | 0.001 |  |
| **Absolute T<sub>reg</sub> counts** |  |  |  |
| Baseline | 5.9±5.0 | 6.5±7.1 | 0.817 |
| Follow-up | 3.6±2.7 | 4.8±2.9 | 0.244 |
| Change from baseline | -2.3 (0.35 to -5.1) | -1.7 (2.1 to -5.3) |  |
| P value | 0.082 | 0.358 |  |
| **Absolute NK Cell count** |  |  |  |
| Baseline | 10.4±9.0 | 9.9±11.4 | 0.898 |
| Follow-up | 17.6±9.5 | 28.3±32.5 | 0.248 |
| Change from baseline | 7.2 (13.0 to 1.3) | 18.3 (38.1 to -1.5) |  |
| P value | 0.020 | 0.067 |  |
| **Neutrophils (%)** |  |  |  |
| Baseline | 63.7±16.6 | 63.9±17.1 | 0.969 |
| Follow-up | 54.4±7.9 | 51.7±9.3 | 0.422 |
| Change from baseline (95% CI) | -9.3 (-1.5 to -17.0) | -12.2 (-2.6 to -21.9) |  |
| P value | 0.023 | 0.017 |  |
| **Absolute neutrophil count** |  |  |  |
| Baseline | 4457.1±4597.1 | 4615.4±2562.2 | 0.914 |
| Follow-up | 3985.7±1346.1 | 4046.1±1158.0 | 0.902 |
| Change from baseline (95% CI) | -3555.4 (1581.4 to -2524.3) | -569.3 (799.3 to -1937.8) |  |
| P value | 0.628 | 0.383 |  |
| **Lymphocyte (%)** |  |  |  |
| Baseline | 25.7±12.6 | 27.2±14.7 | 0.776 |
| Follow-up | 32.8±7.1 | 37.0±7.0 | 0.135 |
| Change from baseline (95% CI) | 7.1 (13.7 to 0.5) | 9.8 (18.5 to 1.1) |  |
| P value | 0.036 | 0.031 |  |
| **Absolute lymphocyte counts** |  |  |  |
| Baseline | 1250.0±462.7 | 1653.8±773.1 | 0.109 |
| Follow-up | 2342.9±734.5 | 2846.1±810.0 | 0.103 |
| Change from baseline (95% CI) | 1092.9 (1515.0 to 670.7) | 1192.3 (1698.8 to 685.8) |  |
| P value | <0.0001 | <0.0001 |  |
| **Th cell response** | **SoC + Placebo** | **SoC + IP** |  |
| **Ab Th1** |  |  |  |
| Baseline | 209.5 (45.3 to 419.8) | 164 (27 to 501) | 0.861 |
| Follow-up | 347 (215.8 to 529.3) | 430 (235 to 683) | 0.338 |
| P value for change | 0.098 | 0.023 |  |
| **Ab Th2** |  |  |  |
| Baseline | 143 (66.5 to 230) | 164 (80 to 398) | 0.318 |
| Follow-up | 424 (343.7 to 593.8) | 593 (351 to 768) | 0.338 |
| P value for change | 0.005 | 0.003 |  |
| **Antibody levels** | **SoC + Placebo** | **SoC + IP** |  |
| **IgG** |  |  |  |
| Baseline | 1301.4±284.9 | 1593.6±309.5 | 0.015 |
| Follow-up | 1132.3±223.9 | 1356.6±260.5 | 0.021 |
| Change from baseline (95% CI) | -169.1 (-42.9 to -295.4) | -237.0 (-79.9 to -394.1) |  |
| P value | p=0.012 | p=0.007 |  |
| **IgM** |  |  |  |
| Baseline | 117.7±77.2 | 98.8±40.6 | 0.438 |
| Follow-up | 113.0±62.9 | 89.9±28.4 | 0.420 |
| Change from baseline (95% CI) | -4.7 (8.2 to -17.5) | -8.9 (8.8 to -26.7) |  |
| P value | p=0.450 | p=0.295 |  |
| **COVID Antibodies** | **Placebo** | **Drug** |  |
|  | 5.1±2.5 | 6.0±2.7 | 0.353 |

### Safety

There were no serious adverse events in the drug arm. One SAE occurred in placebo (+SoC) arm, which was ‘prolongation of hospitalization’ due to Covid related pneumonia, which got resolved later. LFT, RFT were not deranged in any of the patient, confirming safety. No drug-drug interaction with the SoC was seen.

## DISCUSSION

Since December 2019, COVID -19 pandemic has been around the globe affecting millions of individuals with significant contribution in morbidity and mortality. Despite being present for over a year, definite therapies are limited for COVID-19. Modulating immunological response with steroids has been found beneficial in hospitalized patients. However, in a large majority of patients who have mild to moderate disease, effective treatment is necessary to prevent disease progression as well as shedding of the virus to lower the transmission rates. Thus, reducing the viral load may assist in achieving this objective. Viral load independently correlates with the risk of intubation and in-hospital mortality [12,13]. We observed that IP was associated with significant reduction in viral load at day 4. Reducing the viral load can help in early symptom recovery. It was evident as NRC was significantly reduced.

Anti-viral activity of IP therefore can be helpful in early recovery of mild to moderate COVID-19 patients. Among currently employed antiviral agents, early treatment with favipiravir was reported to be associated with lower in VL at day 6 In hospitalized patients with COVID-19 [14].

Another antiviral agent that is used widely is remdesivir. However, remdesivir did not appear to affect rates of viral RNA load decline and mortality when compared with placebo in severe hospitalized COVID-19 patients [15]. Thus, initiating an antiviral agent before the peak viral load might be necessary. It may be difficult in clinical situation to predict the time of peak VL. In this study, IP was initiated as soon as individuals were tested positive for COVID-19 which might have helped in early improvement in clinical NRS score.

In addition to lowering VL, modulating immune response is now an established strategy. In inflammatory response, Th1 pathway is associated with increase in the release of proinflammatory cytokines like IL-1, IL-2, IL-12, TNF-α, IFN-γ, etc. Dysregulated immune response is associated with development of cytokine storm. Most recent data demonstrated that COVID-19 might affect lymphocytes, especially T lymphocytes. The absolute number of T lymphocytes, CD4+ T cells, and CD8+ T cells decreased in infected individuals [16]. Immunomodulatory activity of single plants as well as polyherbal formulations from Ayurveda are well reported [17]. Modulation of immune response was evident in our study. Though placebo with SoC also showed significant effect, the change in Absolute CD3, CD4 and T helper cells was numerically higher with IP treatment. Tregs and their functions are compromised in severe COVID-19 patients, engendering unrestrained immune cell activation resulting in damaged lungs in severe COVID-19 patients [18]. There is less reduction of Tregs in the IP group as compared to the placebo group indicates a good inflammatory control in the IP group. In severe COVID-19 patients, NK cell number and function is reduced, resulting in decreased clearance of infected and activated cells, and unchecked elevation of tissue-damaging inflammation markers. Restoration of NK cell effector functions has the potential to correct the delicate immune balance required to effectively overcome SARS-CoV-2 infection [19,20].Though the statistical significance was not reached, increase in the number of NK cells was higher in IP group than that of placebo. It probably indicates a potential to preserve and enhance the NK cell function that might assist in viral clearance.

The in-vitro study conducted with study drug with remdesivir used as a positive control for viral inhibition. Inhibition of virus replication is determined based on the fold change in the Ct value in TS-treated cells compared to the control. After 24 hours, % inhibition of E-gene was 9.10% and of N gene, it was 21.93%. After 48 hours, N gene inhibition was same as 24 hours however E gene was further inhibited to 28.43% (*data on file*). Thus, clinical effects of viral load reduction identified with qRT-PCR are supported by the in-vitro data.

Protocol of the study was published in BMC’s ‘Trials’ (2020)21:943. It was for the first time that any study protocol pertaining to an Ayurveda intervention was published in the BMC journal. (https://doi.org/10.1186/s13063-020-04906-x)

At individual level, each ingredient has unique properties and for example, Guduchi (*Tinospora cordifolia*), Haritaki (*Terminalia chebula*),Amalaki (*Eblica officinalis*) has shown efficacy as anti-viral drugs, as an anti-oxidant, as an immune modulator; for Amalaki, a paper entitled, ‘Structure-based drug designing for potential antiviral activity of selected natural products from Ayurveda against SARS-CoV-2 spike glycoprotein and its cellular receptor’. At least four natural compounds from *Tinospora cordifolia* showed high binding efficacy against SARS-CoV-2 targets involved in attachment and replication of the virus, hence validating the merit of using *Tinospora cordifolia* in the clinical management of infection caused by SARS-CoV-2. There is a certain efficacy of natural compounds from *Tinospora cordifolia* against SARS-CoV-2 protease, surface glycoprotein and RNA polymerase [21-22].Extract of *T. chebula* increased spleen lymphocyte proliferation. Based on RT-PCR analysis, the expression of cytokines (IL-2, IL-10 and TNF-α), was more in *T. chebula* -treated than in other control groups [23]. Antioxidant properties of *Piper longum, Zinziber officinale* are also well-known [24,25]. Antiviral activity of Vidanga (*Embelia ribes*) against Influenza virus and antioxidant activity are well established [26]. Anti-inflammatory, antiviral effects of Yashtimadhu (*Glycyrrhiza glabra*) are also well-known [27,28].Shatavari (*Asparagus racemosus*) has many activities including antioxidant and Immunomodulatory activities [29,30].Minerals in the drug, Shankha (incinerated conch shell) Bhasma is helpful to tackle gastric symptoms related to the disease and Jasath Bhasma (zinc oxide) has an antiviral activity [31]. Jasath Bhasma is the purified zinc from Ayurvedic process to yield optimum effect of zinc supplementation. Due to antioxidant effects of Zinc, it protects against ROS and RNS. Zinc helps modulate cytokine release and induces proliferation of T cells and helps to maintain skin and mucosal membrane integrity.Zinc has a central role in cellular growth and differentiation of immune cells. It is essential for intracellular binding of tyrosine kinase to T cell receptors, required for T lymphocyte development and activation. Zinc supports Th1 response [32].

When we discuss about the possible mechanism of action, the approach is such that the combination acts ‘as a whole’. There are multiple phyto-ingredients and a lot of chemical entities that act on multiple targets in various systems of the body. By virtue of such entity-rich drug, a milieu is created in the body, which is prepared to take care of the infection at multiple levels, organs and systems. Since it has shown positive results in reducing viral load and immune modulation of TH1 response, the same herbo-mineral formulation can have role for prophylactic action towards the virus. Limitation of the study is smaller number of participants.

Currently the formulation is known as Capsule CoviLyzer™ and Capsule Abayakasthaa plus^R^, and is manufactured by Ishaanav Nutraceuticals Pvt. Ltd.

## CONCLUSION

With COVID-19 pandemic being ongoing in 2021, search for potential antiviral therapies is ongoing. This polyherbal combination drug was identified to provide effective antiviral activity in mild to moderate COVID-19 patients. Potential immunomodulatory effects observed with drug can also assist in preventing inflammatory tissue damage and may help reduce the intensity of systemic inflammatory syndrome. The investigational drug was found to be safe with no serious adverse event and it can be safely given along with Standard of Care to enhance its therapeutic activity in patients with mild to moderate COVID-19 disease for reducing viral load, cytokine storm and clinical recovery. Another study on severely affected Covid patients would be worthwhile to explore of this formulation, along with the standard of care.

## Figure Legends

**Image 1:**
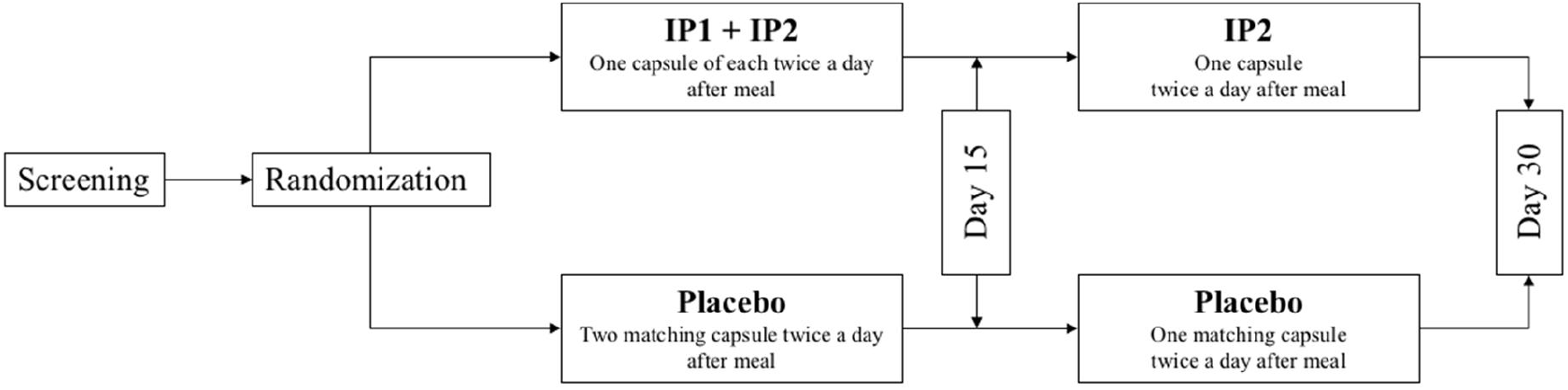
Treatment administration in two groups.

