## Supplementary material for "Efficacy and Safety of Polyherbal formulation as an add-on to the standard of care in mild to moderate COVID-19: A randomized, double-blind, placebo-controlled trial": Protocol--EC Approved

Title of the clinical study: A double blind randomized controlled trial to assess safety and efficacy of Cap. IP in COVID-19 positive patients with mild to moderate severity for early recovery, in restoring respiratory health and in improvement in innate immunity

Protocol number: CoviQuest-01

Protocol version number: 1.2

Date: 1st July 2020

Name of the Investigational Product: Cap. IP (1 and 2) and Placebo

Dosage Form: Capsule

Sponsor: AMAI Trust, Pune

Contract Research Organization: Quest Clinical Services, 14, Samata, Chinchwad, 411033

Principal Investigator: Dr. Kishor Suryawanshi

Co-Investigators: Dr. Pravin Soni, Dr. Suresh Patankar, Dr. Hrishikesh Rangnekar

Sites: YCM Hospital, Pimpri and other TBD

This protocol is written by Quest Clinical Services and is approved by Dr. Suresh Patankar. No part of this shall be published without written permission of Quest Clinical Services or Dr. Patankar.

### Background and Introduction

COVID-19 (previously termed as 2019-nCoV), a novel coronavirus disease with high mortality, emerges as a pandemic disease[i]. In December 2019, a series of acute atypical respiratory disease occurred in Wuhan, China. This rapidly spread from Wuhan to other areas. It was soon discovered that a novel coronavirus was responsible. The novel coronavirus was named as the severe acute respiratory syndrome coronavirus-2 (SARS-CoV-2, 2019-nCoV) due to its high homology (~80%) to SARS-CoV, which caused acute respiratory distress syndrome (ARDS) and high mortality during 2002–2003[ii]

The disease caused by this virus was called Coronavirus disease 19 (COVID-19) and a pandemic was declared by the World Health Organization (WHO). COVID-19 has been impacting a large number of people worldwide, being reported in more than 200 countries and territories.

SARS-CoV-2 virus primarily affects the respiratory system, although other organ systems are also involved. Lower respiratory tract infection related symptoms including fever, dry cough and dyspnea were reported in the initial case series from Wuhan, China[iii]. In addition, headache, dizziness, generalized weakness, vomiting and diarrhea were observed[iv]. It is now widely recognized that respiratory symptoms of COVID-19 are extremely heterogeneous, ranging from minimal symptoms to significant hypoxia with ARDS. In the report from Wuhan mentioned above, the time between the onset of symptoms and the development of ARDS was as short as 9 days, suggesting that the respiratory symptoms could progress rapidly. This disease could be also fatal. A growing number of patients with severe diseases have continued to succumb worldwide. Epidemiological studies have shown that mortalities are higher in elder population[v] and the incidence is much lower in children[vi],[vii].

Current medical management is largely supportive with no targeted therapy available. More treatment modalities are being tested in clinical trials in many countries. A large number of countries have implemented physical distancing and lockdown to mitigate further spread of the virus.

### Symptomatology and Pathophysiology of the disease:

Below table gives an overview of classification of COVID 19 patients:

| TYPE | PRESENTATION |
| --- | --- |
| Asymptomatic | COVID nucleic acid test positive. Without any clinical symptoms and signs and the chest imaging is normal |
| Mild | Symptoms of acute upper respiratory tract infection (fever, fatigue, myalgia, cough, sore throat, runny nose, sneezing) or digestive symptoms (nausea, vomiting, abdominal pain, diarrhea) |
| Moderate | Pneumonia (frequent fever, cough) with no obvious hypoxemia, chest CT with lesions. |
| Severe | Pneumonia with hypoxemia ($SpO2 < 92\%$) |
| Critical | Acute respiratory distress syndrome (ARDS), may have shock, encephalopathy, myocardial injury, heart failure, coagulation dysfunction and acute kidney injury. |

Coronaviruses are enveloped, positive-sense, single-stranded RNA viruses of ~30 kb. They infect a wide variety of host species[viii]. They are largely divided into four genera; α, β, γ, and δ based on their genomic structure. α and β coronaviruses infect only mammals[ix]. Human coronaviruses such as 229E and NL63 are responsible for common cold and croup and belong to α coronavirus. In contrast, SARS-CoV, Middle East respiratory syndrome coronavirus (MERS-CoV) and SARS-CoV-2 are classified to β coronaviruses.

The life cycle of the virus with the host consists of the following 5 steps: attachment, penetration, biosynthesis, maturation and release. Once viruses bind to host receptors (attachment), they enter host cells through endocytosis or membrane fusion (penetration). Once viral contents are released inside the host cells, viral RNA enters the nucleus for replication. Viral mRNA is used to make viral proteins (biosynthesis). Then, new viral particles are made (maturation) and released. Coronaviruses consist of four structural proteins; Spike (S), membrane (M), envelop (E) and nucleocapsid (N)[x]. Spike is composed of a transmembrane trimetric glycoprotein protruding from the viral surface, which determines the diversity of coronaviruses and host tropism. Spike comprises two functional subunits; $S_1$ subunit is responsible for binding to the host cell receptor and $S_2$subunit is for the fusion of the viral and cellular membranes. Angiotensin converting enzyme 2 (ACE2) was identified as a functional receptor for SARS-CoV[xi]. Structural and functional analysis showed that the spike for SARS-CoV-2 also bound to

ACE2. ACE2 expression was high in lung, heart, ileum, kidney and bladder[xii]. In lung, ACE2 was highly expressed on lung epithelial cells.

The symptom of patients infected with SARS-CoV-2 ranges from minimal symptoms to severe respiratory failure with multiple organ failure. On Computerized tomography (CT) scan, the characteristic pulmonary ground glass opacification can be seen even in asymptomatic patients[xiii]. Because ACE2 is highly expressed on the apical side of lung epithelial cells in the alveolar space[xiv],[xv], this virus can likely enter and destroy them. This matches with the fact that the early lung injury was often seen in the distal airway. Epithelial cells, alveolar macrophages and dendritic cells (DCs) are three main components for innate immunity in the airway[xvi]. DCs reside underneath the epithelium. Macrophages are located at the apical side of the epithelium. DCs and macrophages serve as innate immune cells to fight against viruses till adaptive immunity is involved.

Immunological studies were mainly reported in severe COVID-19 patients. Patients with severe diseases showed lymphopenia, particularly the reduction in peripheral blood T cells. Patients with severe diseases were reported to have increased plasma concentrations of proinflammatory cytokines, including interleukin (IL)-6, IL-10, granulocyte-colony stimulating factor (G-CSF), monocyte chemoattractant protein 1 (MCP1), macrophage inflammatory protein (MIP)1α, and tumor necrosis factor (TNF)-α. The more severe conditions patients were in, the higher their IL-6 levels were. $CD4^+$ and $CD8^+$ T cells were activated in those patients as suggested by higher expression of CD69, CD38 and CD44. Higher percentage of checkpoint receptor $Tm3^+PD\text{-}1^+$ subsets in $CD4^+$ and $CD8^+$ T cells showed that T cells were also exhausted. NK group 2 member A (NKG2A), another marker for exhaustion was elevated on CD8+ T cells. Exhaustion of T cells could have led to the progression of the disease. Another interesting finding was that aberrant pathogenic $CD4^+$ T cells with co-expressing interferon (IFN)-γ and granulocyte-macrophage colony-stimulating factor (GM-CSF) were seen in COVID-19 patients with severe disease.[xvii],[xviii]

### Investigational Product:

The IP contains holistic extracts (blend of water and $CO_2$ extracts) of *Shunthi (Zingiber officinale* (Ginger), *Vidanga (Embelia ribes), Yashtimadhu (Glycyrrhiza glabra), Haritaki (Terminalia chebula), Guduchi (Tinospora cordifolia), Shatavari (Asparagus racemosus), Aamalaki (Emblica officinalis), Pippali (Piper longum)* and calcined zinc, *shankha bhasma.*

#### (a) Preclinical experience

There are studies done and literature available on almost each of the component of Cap. IP.

(b) Clinical experience

There are studies done and literature available on almost each of the component of Cap. IP. The dose of each component in the IP is very safe to administer.

**Study Rationale**

COVID -19 is a respiratory illness that has led to a pandemic, affecting more than 200 countries in the world. Due to sudden advent and rapid spread of infection, no antiviral drug has been found to be effective since its onset.

1. While there is no medicine for COVID-19 as of now, enhancing the body's defense system (immunity) plays an important role in maintaining optimum health and the IP (study drug) might possibly play an important role in enhancing immunity.
2. Role of study drug as an antiviral could also be tested.

The proposed trial is to check efficacy of Cap. IP in the management of COVID-19.

We intend to enroll 60 patients which can be extendable to 150 in the study. Eligible and willing patients will either get Cap. IP as an add-on therapy to the SOC (Standard of Care) or placebo (inert substance) as an add-on therapy to the SOC.

Aim: To assess the efficacy and safety of Cap. IP in the management of COVID 19 patients and also in boosting innate immunity

Study Objective(s)

Primary Objective

- To assess efficacy of Cap. IP in boosting innate immunity of patients with COVID-19 infection.

Secondary Objectives

- To assess efficacy of Cap. IP in restoring respiratory health
- To see efficacy of Cap. IP in early recovery of patients and decline in Viral Load
- To assess safety of Cap. IP.

Study Design

Overview of the Study Design

- This is a prospective, blinded, double arm study.
- A total of 60 (Extendable to 150) COVID-19 patients will be enrolled in the study.

- Those fulfilling eligibility criteria and consenting in writing will be enrolled in the study. Considering the very infectious nature of the disease, if EC allows, investigator will sign on patient's behalf initially. Later, once the person is COVID 19 negative, consenting will be done.

- All the patients enrolled in the study will get the Investigational Product (IP-1 and IP-2), or 2 capsule of Placebo two times daily for the 1st 15 days. From 16th day onwards patients will get one capsule twice daily i.e. IP-2 or capsule placebo up to day 30.

- Patients will undergo laboratory investigations upon enrolment and on last visit. RTPCR for COVID will be done on day 5 (± 1 day).

- Patients and doctors treating will not be knowing what the participating subject has received. Subjects will be given coded bottles, containing dose for 30 days.

- No medicine which his/her physician is giving will be stopped. No modifications shall be suggested in their diet and daily routine physical activities.

- Thus, patients enrolled in the study will get the IP / Placebo in addition to the standard of care which they were already receiving at the time of enrolment.

- Half of patients will be getting Cap. IP and half will be getting Cap. Placebo throughout the study duration of 1 month.

Study period for enrolled patients will be 30 days:

- Enrollment (day 0): After Informed Consent Process, Demographics and other information for CRF will be recorded. Investigations: COVID 19 RTPCR, Lymphocyte subset analysis (TH1, TH2, Th17, IL6, NK Cells and CD markers); Immunoglobulin IGG (Serum); Immunoglobulin IGM (Serum); Liver Function Test and Kidney Function Test will be done. Cap. bottle will be given to the patients with the dose lasting for 30 days.
- Day 3±1: COVID-19 RT PCR (quantitative) test will also be conducted on day 3 (±1) on 5 subjects of each group

- Follow-up 1: Day (5±1): COVID-19 RT PCR (quantitative) test will be conducted for all subjects on Day (5±1). CRF will be filled for knowing the overall status.

- Follow up 2 (15th day± 2 days): This follow up will be clinical/telephonic. CRF will be filled for knowing the overall status.

- Follow up 3 (30th day± 2 days): Patients will be followed up at day 30. This visit will be done telephonically. Patient will not visit site but blood investigations shall be carried out. Investigations will be done outside the site, considering that the site is COVID specific hospital. The investigations are; Lymphocyte subset analysis (TH1, TH2, Th17, IL6, NK Cells and CD markers); Immunoglobulin IGG (Serum); Immunoglobulin IGM

(Serum); Liver Function Test and Kidney Function Test will be done. All the unused IP will be collected.

- Sponsor will provide 10 codes (5 from each, IP and placebo, not mentioning which is placebo and IP). These will get the RT PCR for COVID-19 done on 3±1in addition to day (5±1). This will be done to know the graph of viral load. Sr. Ferritin, CRP and D- Dimer will be done for same patients on day 0 and day 30.

Every time subject coming to the scheduled follow-up should be asked to bring the remaining capsules with the bottle.

### Flow chart

Screening

(I/E Criteria, ICF reading)

↓

Enrollment (Day 0)

(Eligibility criteria, Consenting, Clinical Exam, Investigations, QOL,IP Dispense)

↓

COVID-19 RT PCR (quantitative) test on day 3 (±1) for 5 subjects of each group

↓

1st Follow-up: Day (5±1)

COVID-19 RT PCR (quantitative)

↓

2nd Follow-Up: Day (15±2)

[Clinical/telephonic]

↓

3rd Follow-Up: Day (30±2)

[Tele follow-up Blood Investigations at lab]

**Methods and Procedures**

Patients attending COVID set-ups (YCM Hospital, Pimpri, Pune) of the investigators will be screened for their participation in the study. Patients should be COVID-19 positive, diagnosed recently. No discretion for enrollment will be made based on gender, color, religion and cast. Information about the study will be shared with the patients and their eligibility will be checked during screening. Those fulfilling this requirement and who are willing to participate will be given informed consent document and patient information sheet for reading. Adequate time will be given for reading the same and the same will be explained verbally. All questions will be answered to the patients' satisfaction. Patient will be considered as enrolled only after consenting procedure is done.

Considering the highly infectious nature of the disease under consideration, if EC approves, consenting will be done once the patient is COVID 19 negative.

Patients will be counseled about administration of IP (dose, timing, method etc.).

Follow-Up:
After enrollment (Day 0), there will be one clinical and the next two: either telephonic or clinical follow-ups: first follow-up visit at day 5 (±1 day), second follow-up visit at day 15 (±2 days) and third follow-up visit at day 30 (±2 days).

Patients will need to make sure, there is no gap in consuming the IP.

Patients can call the study coordinator anytime for the queries if any.

Unscheduled Visits:

Patients will continue to visit his/her physician at the site for medical care with prior appointments for routine and regular check-ups. Study coordinator will try and combine their study follow-up visit with their regular visit at the site. Patients should, as far as possible, visit the site in case of medical emergencies.

Study Design:

This is a two arm, prospective study to determine safety and efficacy of Cap. IP in COVID-19 positive patients.

After enrollment, they will get the IP / Placebo in addition to their regular treatment they might be receiving for their condition. (2 capsules twice a day for prior 15 days and 1 capsule twice a day from day 16)

A total of 60 patients (Can be extendable to 150) will be enrolled in the study in different groups. The reason behind this sample size that, we can apply paired t test with adequate power for an exploratory study.

**Sample size: Cross- sectional, Cohort and Randomized Clinical Trials**

| | |
|---|---|
| Two sided significance level ( 1 alpha) | 95 |
| Power ( 1-beta. % chance of detecting) | 80 |
| Ratio of sample size, Unexposed/ exposed | 1 |
| % of Unexposed with outcome | 5 |
| % of Exposed with outcome | 35 |
| Odds ratio | 10 |
| Risk/ Prevalence ratio | 7 |
| Risk/ Prevalence | 30 |

| | Kelsey | Fleiss |
|---|---|---|
| Sample Size Exposed | 28 | 27 |
| Sample Size Non exposed | 28 | 27 |
| **Total Sample Size** | **56** | **54** |

**References:**

Kelsey et al, Methods in Observational Epidemiology, 2nd edition, table 12-15, Fleiss, Statistical Methods For Rates and Proportions, formulas 3.18 and 3.19.

**Results are rounded up to the nearest integer**

**Results from OpenEpi, Version 3, open source calculator –SSC cohort.**

Considering drop-outs, 30 in each arm are considered.

Study Population:

COVID-19 positive subjects.

Subject Eligibility:

### (a) Inclusion Criteria

1. Age > 18 years
2. All sexes
3. Case definitions for inclusion in the study will include mild to moderately severe cases as defined in the Guidance on appropriate treatment of suspect/confirmed case of COVI19 issued by Ministry of Health and Family Welfare, Govt of India on 07 Apr 2020.
   Mild: Cases presenting with fever and/or upper respiratory tract illness (Influenza like Illness, ILI); Moderate: Pneumonia with no signs of severe disease (Respiratory Rate 15 to 30/minute, SpO2 90%-94%). OR
4. Laboratory confirmed SARS CoV-2 infection within last 10 days or SARS CoV-2 test result pending with a high clinical suspicion as defined by: Cough of more than 10 days duration OR
5. Bilateral pulmonary infiltrates on chest X Ray / CT scan or new hypoxaemia defined as SpO2 <94% on room air, where no alternative explanation for respiratory symptoms can be given

### (b) Exclusion Criteria

1. Pregnant or lactating women
2. Symptoms of acute respiratory tract infection for more than seven days
3. More than 48 hours have elapsed between meeting inclusion criteria and enrolment
4. Subject is also a participant of any other clinical trial
5. Serious / long-standing co-morbid conditions

**Study Assessments:**

Participants will have a minimum of two follow-ups. Clinical assessment will be done; vitals will be noted, questionnaire will be filled-in and blood will be collected for various parameters.

**Study Conduct**:

Patients screened, eligible and willing will be enrolled in the study and IP will be dispensed. During screening, inclusion-exclusion criteria will be applied. Those fulfilling the criteria will be given detailed information about the study. Of those who are willing to participate will be asked to sign the informed consent form. Study coordinator with the principal or co-principal investigator will participate in the consent process.

Day of consenting will be done on the day of screening. After signing the consent form, on enrollment day, physical examinations will be done; blood will be withdrawn for investigations, case record form will be filled and IP will be dispensed. A copy of duly signed informed consent form will be given to them.

Follow-up Visit 1 [5$^{th}$ day (± 1) of the day of enrollment]: Questions will be asked about adherence, missed doses if any and adverse events if any. NP swab will be collected for RT-PCR of COVID-19

Follow-up Visit 2 [15$^{th}$ day (± 2) of the day of enrollment]: All the procedures for this visit will be the same as that of Visit 1, except that swab will not be collected for RT PCR.

Follow-up Visit 3 [30$^{th}$ day (± 2) of the day of enrollment]: All the procedures for this visit will be the same as that of Visit 2, and in addition, blood will be drawn for various markers.

Discontinued Patients:

Patients will be withdrawn/dropped from the study if they violate the protocol e.g. by participating in other study, by not taking the IP, not coming to the hospital for follow-up etc. Patients developing any serious disease, patients wanting to withdraw will also be withdrawn from the study.

Patients who have not turned for clinical follow-up will be called telephonically up to three times after the window period is over. During consenting they will be asked if it is alright with them.

However, as it is a 'intent-to-treat' (ITT) study design, data collected of such patients, till they are dropped out/withdrawn from the study will be included in the analysis.

Patients will be insisted to visit during the window-period. Those who miss this will still be continued into the study. On case-to-case basis, protocol waivers will be granted by the co-PI and/or the PI.

Protocol violations will be notified to the EC and to the concerned person responsible for such violations. Appropriate training will be given by the sponsor or by Quest clinical services. Quest clinical services have rights to change the person and notify the EC accordingly.

In case of continuous or repetitive violations hampering the study design in total or jeopardizing the safety of patients, study will be terminated after informing the EC.

Randomization Process:
As this is a double-blind study, and with alphanumeric coded set of IP/placebo, subjects will be automatically randomized to two groups. Blind will be broken after the last follow-up of last enrolled subject is over.

**Study Treatment:**

(a) Dosing schedule (dose, frequency, and duration):

Participants will be dispensed IP, which is Cap IP or Cap. Placebo.

Dose of the IP is: For 15 days two Capsules two times a day, and from 16$^{th}$ day one capsule two times a day upto day 30. It is to be administered orally with plain water.

IP, in any dose, needs to be taken for a period of 30 days in total; from the day of enrollment.

IP will be given from the study site by the study coordinator, free of cost. All other concomitant medicines prescribed by the treating physician/doctor has to be taken by the patient.

Half of the patients enrolled will get Placebo (inert substance) in the capsule throughout the study period. However, neither patient nor the investigators will know, who is receiving what.

(b) Study drug supplies and administration: Study medication will be provided by AMAI, Pune, which is the sponsor of the trial. Quest clinical services will distribute the same to the enrolled patients in the manner mentioned in this protocol. Investigational drug formulation has been manufactured following all the regulations.

The IP will be securely stored in cupboards; at room temperature. Expiry date will be verified before giving the IP to the patients. CRC will document the code on the bottle to given to corresponding patient.

(c) Dose modification for study drug: In case of any serious adverse event, that warrants to break the blind, sponsor will be contacted. Dose might be modified or IP might be stopped, in the best safety interest of the patient.

(d) Possible drug interactions: As this is an Ayurvedic compound, involving safe profile herbs, no drug interactions are expected.

(e) Concomitant therapy: As a part of the study design, all the medicines that the patient is supposed to take for his/her other condition(s), patient might be having, will be allowed. There is no drug that is not allowed, provided, it is prescribed by the treating physician. If the PI or co-PI decides that by remaining in the study could be harmful for his/her health, patient will be discontinued from the study.

(f) Blinding procedures:

Placebo is used in this study, to allow blinding of investigators and subjects to treatment and minimize bias in the evaluation of safety and efficacy assessments,

Subjects, investigator staff and personnel performing the study assessments will remain blinded to the identity of the treatment from the time of randomization. Monitoring will be done by unblinded personnel from sponsor.

Following measures will be applied to keep required personnel blinded.

1. Sponsor will provide coded bottles containing IP or placebo in those.
2. The list of which bottle contains what will not be shared with any site member or with the CRO.

Adverse Events:

Very serious allergic reaction to this drug should be rare. However, patient will be asked to seek immediate medical attention if he/she notices any symptoms of a serious allergic reaction, including: rash, itching/swelling, severe dizziness, trouble breathing.

Of particular interest, these drugs are generally regarded as safe.

Reporting of the adverse events will be done in following format:

1. Patient Details:
   Initials and other relevant identifier (PID)*
   Gender:
   Age:
   Weight:
   Height:
2. Suspected Drug(s)
   Generic name of the drug(s)*
   Indication(s) for which suspect drug was prescribed or tested
   Dosage form and strength
   Daily Dose and Regimen
   Route of administration
   Starting date and time of day
   Stopping date and time of day or duration of treatment
3. Other Treatment(s)
   The same information for concomitant drugs (including non-prescription/OTC drugs) and non-drug therapies, as for the suspected drug(s) will be provided.
4. Details of Suspected Adverse Drug Reaction(s)
   Full description of reaction(s) including body site and severity, as well as the criterion (or criteria) for regarding the report as serious will be provided. In addition to a description of the reported signs and symptoms, whenever possible, a specific diagnosis for the reaction will be described.*

Start date (and time) of onset of reaction

Stop date (and time) or duration of reaction

De-challenge and re-challenge information

Setting (e.g., hospital, out-patient clinic, home, nursing home)

5. Outcome

   Primary Outcome:

   Efficacy of Cap. IP in COVID 19 positive patients.

   Secondary Outcomes:

   Information on recovery and any sequelae; results of specific tests and/or treatment that may have been conducted will be provided. For a fatal outcome, cause of death and a comment on its possible relationship to the suspected reaction; any post-mortem findings will be included. Other information: anything relevant to facilitate assessment of the case, such as medical history including allergy, drug or alcohol abuse; family history; findings from special investigations etc. will also be included.

   Test of batteries will throw light on the change in innate immunity of the participants.

6. Details about the Investigator*

   Name

   Address

   Telephone number

   Profession (specialty)

   Date of reporting the event to Ethics Committee:

   Signature of the Investigator

   Information marked '*' must be provided.

**Ethical Considerations**

(a) Risk/benefit assessment:

From the data available of the product, there are no major health risks to the participants. However, if investigators find any information that might be associated with the IP, it will be conveyed to all the participants.

(b) Ethics Committee review and communications:

Essential documents will be submitted to the Institutional / Independent Ethics Committee (IEC) for its review. No patient will be enrolled in the study unless the IEC approves the study.

Updates will be given to the IEC each month. Serious adverse events will be reported within 24 hours of their knowledge.

All the communication with the IEC will be filed separately. Quest clinical services will communicate with the IEC on behalf of the sponsor.

(c) Informed consent process:

Information about the IPs and about the trial, procedures of the trial and risks-benefits will be conveyed to the patient so that s/he can take an informed decision. Patient Information Sheet (PIS) and Informed Consent Form (ICF) will be given to patients for reading. Illiterate patients will be read out in presence of an Impartial Witness. All questions and doubts of the patients will be resolved. Only eligible patients, fulfilling inclusion-exclusion criteria and willing to participate will be enrolled only through informed consent process. Each enrolled patient will get a unique identification number. Study coordinator will help PI and/or co-PI in conducting this process. In view of the contagious nature of the disease, if EC allows, consenting will be done post COVID-19 negative result, and s/he will be conveyed orally by the investigator, of the study.

(d) Subject confidentiality; Ownership of data and Coding procedures: Name of the patient will not be mentioned in the case record form or in the reports. There will be a mention of Patient Identification Number (PID) and initials of the patient instead. Sheet linking this PID and name of the patient will be safely secured with the study coordinator. Only members of IEC, monitors, auditors, regulatory persons will have access to the data of the enrolled patients. In all such cases, confidentiality about the participant's identity will be maintained.

In case such as managing adverse event, identity will be revealed only to the concerned.

Coding will be done by the study coordinator. Participants' identity will be concealed by a number. Study coordinator will keep the file linking these numbers to names in a safe and secured place at the site. Un-blinding list of the IP will be with the sponsor, and will not be shared with the team involved in the site activities.

Data, Study Monitoring and Supervision:

Data of the participants will be filled in by the study coordinator. Co-PI and PI will have rights to correct the data whenever needed. All the procedures will be carried out by adhering to the Good Clinical Practices (GCP). Monitor will have access to the study documents. Sponsor of the study can make audit of the study with prior appointment with the PI and the study coordinator.

**Investigational Product Management:**
(a) Investigational product packaging:
Capsules will be dispensed in the coded and sealed containers, which will last for 30 days.

(b) Dosing required during the study:

Total weight of each capsule in both the arms will be the same.

Dose is: two capsules twice a day for 15 days and from 16$^{th}$ day one capsule twice a day, preferably empty stomach. One of the two capsules given during 1$^{st}$ 15 days contains herbs to increase appetite and to digest the undigested. (*Deepan-Pachan).*

Timings should be: IP-1: before both meals, and IP-2 after 30 minutes after both meals

(c) Method of packaging, labeling, and blinding of study substances:

IP / Placebo, will be given to the enrolled patients. Labeling of the pouches/bottles will have all the essentials required. Blinding and coding will be done by the sponsor.

(d) Method of assigning treatments to Subjects and the Subject identification code numbering system:

Alfa-numeric identification numbers will be assigned to the participants.

(e) Storage conditions for study substances:

IP will be securely stored in cupboards; at room temperature.

(f) Investigational product accountability:

Bottles of IPs in sufficient quantity for a period of 30 days for each participant. Receipt and dispensing book will be maintained for the IP.

Participants will be asked to return, partially completed or unopened bottles. Proper recording will be maintained for dispensing and retrieval of the IP.

(g)Policy and procedure for handling unused investigational products:

All the unused IP will be sent to the sponsor.

**Data Analysis**

Blinding will be broken at 30 days i.e.at the end of the study.
Paired t test and any other appropriate test will be applied for analysis.
As this is intent-to-treat design, all the data, even the dropped-out patients' and lost to follow-up patients' collected data will be analyzed.

**Study Assessment Table**

| Visit→ | Screening | Visit Enrolment | 1st Follow up | 2nd Follow up | 3rd Follow up |
|---|---|---|---|---|---|
| Study Procedure | (Day -2 to Day 0) | Day 0 | Day 5th (±1) | Day 15th (±2) | Day 30th (±2) |
| ICF | X | X | | | |
| I/E Criteria | X | X | | | |
| Demographics | X | | | | |
| Vitals and Physical Examination | | X | X | X | X |
| Medical History | X | | | | |
| Questionnaire | | X | X | X | X |
| Investigations | | X*# | X# | | X* |
| Bottle Dispensing | | X | | | |
| IP / Placebo Accountability | | | X | X | X |
| AE/SAE | X | X | X | X | X |

Investigations –

*TH1, TH2, Th17, IL6, NK Cells and CD markers; Immunoglobulin IGG (Serum); Immunoglobulin IGM (Serum); Liver Function Test and Kidney Function Test

Around 2 ml of blood will be stored for additional biomarker in Immunology and Virology.

#COVID-19 RT PCR (quantitative): This test will be conducted on day 3±1 day on 5 subjects of each group. Same group: Sr. Ferritin, CRP and D-Dimer will be done on day 0 and 5.

Clinical investigations like chest X ray, ECG will be done as per physician's discretion. Vitals and Physical Examination: If physician is not in a position to perform, s/he will extrapolate the data from the available information.

**References**

---

[i]World Health Organization. Coronavirus disease 2019 (COVID-19) Situation Report – 73 [Internet]. 2020 [cited 2020 May 9]. Available from: https://www.who.int/emergencies/diseases/novel-coronavirus-2019/situation-reports

[ii]Ksiazek T.G., Erdman D., Goldsmith C.S., Zaki S.R., Peret T., Emery S., Tong S., Urbani C., Comer J.A., Lim W., Rollin P.E., Dowell S.F., Ling A.E., Humphrey C.D., Shieh W.J., Guarner J., Paddock C.D., Rota P., Fields B., DeRisi J., Yang J.Y., Cox N., Hughes J.M., LeDuc J.W., Bellini W.J., Anderson L.J., SW Group A novel coronavirus associated with severe acute respiratory syndrome. N Engl J Med. 2003;348:1953–1966. [PubMed] [Google Scholar]

[iii]Huang C., Wang Y., Li X., Ren L., Zhao J., Hu Y., Zhang L., Fan G., Xu J., Gu X., Cheng Z., Yu T., Xia J., Wei Y., Wu W., Xie X., Yin W., Li H., Liu M., Xiao Y., Gao H., Guo L., Xie J., Wang G., Jiang R., Gao Z., Jin Q., Wang J., Cao B. Clinical features of patients infected with 2019 novel coronavirus in Wuhan, China. Lancet. 2020;395:497–506.[PMC free article] [PubMed] [Google Scholar]

[iv]Shi H., Han X., Jiang N., Cao Y., Alwalid O., Gu J., Fan Y., Zheng C. Radiological findings from 81 patients with COVID-19 pneumonia in Wuhan, China: a descriptive study. Lancet Infect Dis. 2020;20:425–434. [PMC free article] [PubMed] [Google Scholar]

[v]Zhou F., Yu T., Du R., Fan G., Liu Y., Liu Z., Xiang J., Wang Y., Song B., Gu X., Guan L., Wei Y., Li H., Wu X., Xu J., Tu S., Zhang Y., Chen H., Cao B. Clinical course and risk factors for mortality of adult inpatients with COVID-19 in Wuhan, China: a retrospective cohort study. Lancet. 2020;395:1054–1062. [PubMed] [Google Scholar]

[vi]Lu X., Zhang L., Du H., Zhang J., Li Y.Y., Qu J., Zhang W., Wang Y., Bao S., Li Y., Wu C., Liu H., Liu D., Shao J., Peng X., Yang Y., Liu Z., Xiang Y., Zhang F., Silva R.M., Pinkerton K.E., Shen K., Xiao H., Xu S., Wong G.W.K., T Chinese Pediatric Novel Coronavirus Study SARS-CoV-2 infection in children. N Engl J Med. 2020 doi: 10.1056/NEJMc2005073. [PMC free article] [PubMed] [CrossRef] [Google Scholar]

[vii]Qiu H., Wu J., Hong L., Luo Y., Song Q., Chen D. Clinical and epidemiological features of 36 children with coronavirus disease 2019 (COVID-19) in Zhejiang, China: an observational cohort study. Lancet Infect Dis. 2020 doi: 10.1016/S1473-3099(20)30198-5.[PMC free article] [PubMed] [CrossRef] [Google Scholar]

[viii]Channappanavar R., Zhao J., Perlman S. T cell-mediated immune response to respiratory coronaviruses. Journal. 2014;59:118–128. [PMC free article] [PubMed] [Google Scholar]

[ix]Rabi F.A., Al Zoubi M.S., Kasasbeh G.A., Salameh D.M., Al-Nasser A.D. SARS-CoV-2 and Coronavirus disease 2019: what we know so far. Journal. 2020;9 [PMC free article][PubMed] [Google Scholar]

[x]Bosch B.J., van der Zee R., de Haan C.A., Rottier P.J. The coronavirus spike protein is a class I virus fusion protein: structural and functional characterization of the fusion core complex. Journal. 2003;77:8801–8811. [PMC free article] [PubMed] [Google Scholar]

[xi]Li W., Moore M.J., Vasilieva N., Sui J., Wong S.K., Berne M.A., Somasundaran M., Sullivan J.L., Luzuriaga K., Greenough T.C., Choe H., Farzan M. Angiotensin-converting enzyme 2 is a functional receptor for the SARS coronavirus. Journal. 2003;426:450–454.[PMC free article] [PubMed] [Google Scholar]

[xii]Zou X., Chen K., Zou J., Han P., Hao J., Han Z. Single-cell RNA-seq data analysis on the receptor ACE2 expression reveals the potential risk of different human organs vulnerable to 2019-nCoV infection. Journal. 2020 doi: 10.1007/s11684-020-0754-0.[PMC free article] [PubMed] [CrossRef] [Google Scholar]

[xiii]Guan W.J., Ni Z.Y., Hu Y., Liang W.H., Ou C.Q., He J.X., Liu L., Shan H., Lei C.L., Hui D.S.C., Du B., Li L.J., Zeng G., Yuen K.Y., Chen R.C., Tang C.L., Wang T., Chen P.Y., Xiang J., Li S.Y., Wang J.L., Liang Z.J., Peng Y.X., Wei L., Liu Y., Hu Y.H., Peng P., Wang J.M., Liu

J.Y., Chen Z., Li G., Zheng Z.J., Qiu S.Q., Luo J., Ye C.J., Zhu S.Y., Zhong N.S., C China Medical Treatment Expert Group for Clinical characteristics of coronavirus disease 2019 in China. Journal. 2020 doi: 10.1056/NEJMoa2002032. [PMC free article] [PubMed] [CrossRef] [Google Scholar]

[xiv]Hamming I., Timens W., Bulthuis M.L., Lely A.T., Navis G., van Goor H. Tissue distribution of ACE2 protein, the functional receptor for SARS coronavirus. A first step in understanding SARS pathogenesis. Journal. 2004;203:631–637. [PMC free article][PubMed] [Google Scholar]

[xv]Jia H.P., Look D.C., Shi L., Hickey M., Pewe L., Netland J., Farzan M., Wohlford-Lenane C., Perlman S., McCray P.B., Jr. ACE2 receptor expression and severe acute respiratory syndrome coronavirus infection depend on differentiation of human airway epithelia. Journal. 2005;79:14614–14621. [PMC free article] [PubMed] [Google Scholar]

[xvi]Yoshikawa T., Hill T., Li K., Peters C.J., Tseng C.T. Severe acute respiratory syndrome (SARS) coronavirus-induced lung epithelial cytokines exacerbate SARS pathogenesis by modulating intrinsic functions of monocyte-derived macrophages and dendritic cells. Journal. 2009;83:3039–3048. [PMC free article] [PubMed] [Google Scholar]

[xvii] Zhou Y., Fu B., Zheng X., Wnag D., Zhao C., Qi Y., Sun R., Tian Z., Xu X., Wei H. Pathogenic T cells and inflammatory monocytes incite inflammatory storm in severe COVID-19 patients. Journal. 2020 [Google Scholar]

[xviii]Qin C., Zhou L., Hu Z., Zhang S., Yang S., Tao Y., Xie C., Ma K., Shang K., Wang W., Tian D.S. Dysregulation of immune response in patients with COVID-19 in Wuhan, China. Journal. 2020 doi: 10.1093/cid/ciaa248. [PMC free article] [PubMed] [CrossRef] [Google Scholar]
