## Supplementary material for "Efficacy and Safety of Polyherbal formulation as an add-on to the standard of care in mild to moderate COVID-19: A randomized, double-blind, placebo-controlled trial": ETHICS STATEMENT

### Pimpri Chinchwad Municipal Corporation's
### Y.C.M. Hospital, Pimpri - 411018

**Approval Letter of PCMC's PGI YCMH Ethics Committee (Registration No. ECR/1236/Inst/MH/2019)**

YCMH/WS/5/KV/705/2020
Dt- 12/6/2020

**To,**
**Dr. Kishor H. Suryawanshi,**
**Principal Investigator, PGI YCMH.**
**Ref: IEC code - DT-05/01-2020**
**Project title: Study/Protocol No. - CoviQuest-01**

**Dear Dr. Kishor H. Suryawanshi,**
Institutional Ethics Committee reviewed and discussed your application (dated) to conduct the research study entitled – **"A double blind randomized controlled trial to assess safety and efficacy of Cap. IP in COVID-19 positive patients with mild to moderate severity for early recovery, in restoring respiratory health and in improvement in innate immunity"** during the IEC meeting held on 09th June 2020.

The following documents were reviewed and approved:
1. Project Submission form (IEC Proforma).
2. **Study protocol** (including protocol amendments), **dated 01.06.2020, Version 1.1**
3. **Research committee**/department/funding agency/doctoral committee/scientific committee **approval, dated 02nd June, 2020.**
4. **Case Recording Form** and **Informed Consent Document** in English and Marathi, **Version 1.1, dated 01.06.2020,**
5. **DCGI Approval** Letter – Not Applicable.
6. **Curriculum vitae** of a Principal investigator and Co Principal Investigator,
7. **Quote** of **Insurance policy**/compensation for participation and for serious adverse events occurring during the study participation – **Dated** .
8. **Investigator's Agreement** with the sponsor,
9. **Investigator's undertaking**, Dated **04.06.2020,**
10. **Clinical Trial Agreement** (CTA)/Memorandum of Understanding (MOU)/Material Transfer Agreement (MTA), if applicable – **Dated - June, 2020,**
11. **Clinical Trials Registry-India (CTRI),** in case of drug trial require at time of submission but in other case this must be done after approval of the study but before initiation.
12. **Investigator's and Information Brochure - dated ----, Version ---,**
13. **Confidentiality & Conflicts of Interest** Document, Dated 04.06.2020,

**This decision was taken by consensus.**
**Neither PI nor any of proposed study team members participated during the decision making of the IEC.**

#### Pimpri Chinchwad Municipal Corporation's
#### Y.C.M. Hospital, Pimpri - 411018

The following members of the Institutional Ethics committee (IEC) were present at the meeting held on Date 09.06.2020

**Place**: - Seminar Hall, 2nd Floor, PCMC's PGI, Y.C.M. Hospital, Pimpri, Pune – 411018, Maharashtra.

**Time – 11 – 2 Pm**.

| Sr. No. | Name of member | Position on IEC | Affiliation | Gender |
|---|---|---|---|---|
| 1. | Dr. Abhijeet Tilak | Chairman | Not Affiliated | Male |
| 2. | Dr. Tushar Patil | Member Secretary | Affiliated | Male |
| 3. | Dr. Sanjay Juvekar | Social Scientist | Not Affiliated | Male |
| 4. | Dr. Maroti Gaikwad | Clinician | Affiliated | Male |
| 5. | Dr. Deepali Ambike | Clinician | Affiliated | Female |
| 6. | Mr. Chandrakant Indalkar | Legal Expert | Not Affiliated | Male |
| 7. | Dr. Mangal Supe | Clinician | Affiliated | Female |
| 8. | Mr. Dnyaneshwar Shinde | Lay Person | Not Affiliated | Male |
| 9. | Dr. Pravin N. Soni | Clinician | Affiliated | Male |
| 10. | Dr. Vaishali Bhamre | Independent Consultant, Expert | Not Affiliated | Female |

The trial is approved in its presented form. The **approval is valid until one year or duration of project whichever is later from the date of sanction**. You may make a written request for renewal / extension of the validity, along with the submission of annual status report.

**Following points must be noted: -**

1. **IEC should be informed of the date of commencement of study** (AN5-V1/IEC PGI YCMH SOP 06/V1) and annual progress.
2. IEC has approved recruitment of 150 patients on this study; Number may vary, as it's a multicenter study.
3. PI and other investigators should co-operate with IEC, which may monitor the trial from time to time.
4. The decision was arrived at through consensus. **Neither PI nor any of proposed study team members was present during the decision making of the IEC.**
5. At the time of PI's retirement/intention to leave the institute, study responsibility should be transferred to a colleague after obtaining clearance from HOD and getting IEC concurrence and submitting status report, including accounts details should be submitted to HOD, IEC and extramural sponsors.
6. The IEC functions in accordance with the GCP-CDSCO/ICMR/Schedule Y guidelines/ICH-GCP, Clinical Trial Rules 2019.

7. **New information or any AE/SAE**, which could affect any study, **must be communicated** to **IEC and sponsors**. The PI should report SAEs occurred for IEC approved studies within 7 days of the occurrence of the SAE. **If the SAE is 'Death'**, the **Bioethics cell should receive** the **SAE reporting form within 24 hours** of the occurrence.

8. **In the events of any protocol amendments**, IEC must be informed and the amendments should be highlighted in clear terms as follows:

**a.** The exact alteration/amendment should be specified and indicated where the amendment occurred in the original project. (Page no. Clause no. etc.)

**b.** The PI must comment how proposed amendment will affect the ongoing trial.

**c.** Alteration in the budgetary status, staff requirement should be clearly indicated and the revised budget form should be submitted.

**d.** If the amendments require a change in the consent form, the copy of revised Consent Form should be submitted to Ethics Committee for approval.

**e.** If the amendment demands a re-look at the toxicity or side effects to patients, the same should be documented.

**f.** If there are any amendments in the trial design, these must be incorporated in the protocol, and other study documents. These revised documents should be submitted for approval of the IEC, only then can they be implemented.

**g.** Approval for amendment changes must be obtained prior to implementation of changes. The amendment is unlikely to be approved by the IEC unless all the above information is provided.

9. Any deviation/violation/waiver in the protocol must be informed to the IEC as detailed in IEC PGI YCMH SOP 09/V1.

10. If project/drug/device trial initiation not done in next 6 months from date of approval from IEC, further extension will not be granted and it will require resubmission to IEC.

11. The PI, CO PI needs to be trained in Good Clinical Practice Guidelines.

Thanking You,

Yours Sincerely,

**Signature of the Member Secretary**

Date 12·06·2020

Name – Dr. Tushar V. Patil.

Member Secretary, IEC,
PCMC's PGI, YCMH
ECR/1236/Inst/MH/2019

**Signature of the Chairperson**

Date 12/06/2020

Name – Dr. Abhijeet V. Tilak.

Chairman, IEC,
PCMC's PGI, YCMH
ECR/1236/Inst/MH/2019
